# Low back pain care pathways – is the last provider seen more important than the first: A retrospective cohort study

**DOI:** 10.1101/2022.10.27.22281624

**Authors:** David Elton, Meng Zhang

## Abstract

**Background:** The associations between the type of health care provider (HCP) initially contacted by an individual with low back pain (LBP), guideline concordance, and cost of care are well understood. Less is known about types of HCP that become involved after the initial HCP, particularly the last HCP seen. The purpose of this retrospective cohort study was to examine sequence patterns in the types of HCP involved in an episode of LBP, exposure to 13 types of services, and total cost.

**Methods:** A US national sample of commercially insured individuals with a single episode of non-surgical LBP occurring in 2017-2019 was analyzed using episode of care unit of analysis. The primary independent variable was the type of initial contact HCP. Dependent measures included types of subsequent HCPs involved in an episode of LBP, rate of use of 13 types of health care services, and total cost.

**Results:** 503,958 individuals aged 18 years and older initially contacted 196,522 different HCPs for an episode of non-surgical LBP. 30.2% of individuals saw a second HCP, 11.8% a third, 5.2% a fourth and 2.5% a fifth. Compared to primary care providers (PCP) (71.0% of episodes), when chiropractors (DC) were the initial contact HCP they were the most likely to be the only and last HCP seen (84.0% of episodes) (risk ratio 1.18, 95% confidence interval 1.18-1.19). DCs were also the most likely to be the last HCP seen when the second HCP in an episode (72.7%) (1.10, 1.09-1.12), the third HCP (67.7%) (1.10, 1.08-1.13), or the fourth HCP (61.6%) (1.13, 1.08-1.17). Physical therapists (PT), while less likely than PCPs to be the only and last HCP seen (60.0%) (0.84, 0.83-0.86), were more likely than PCPs to be the last HCP when the second (70.9%) (1.08, 1.06-1.09), third (63.2%) (1.03, 1.00-1.06), or fourth HCP (60.7%) (1.11, 1.06-1.16) seen during an episode. PTs were the only type of HCP involved in more episodes as the second HCP (7,344) than as the initial HCP (5,114).

**Conclusions:** DCs are the most likely to be the last HCP seen for an episode of non-surgical LBP. PTs are as likely as DCs to be the last HCP seen except when initially contacted by an individual with LBP. There are numerous potential confounders and limitations to consider when interpreting this novel finding.

## Background

Low back pain (LBP) clinical practice guidelines (CPG) recommend a stepped approach to management with an emphasis on non-pharmaceutical and non-interventional first-line approaches for most LBP.^1,2^ The type of health care provider (HCP) initially contacted by an individual with LBP, and associated CPG concordance and cost, is the subject of a growing body of research.^3-8^ A high proportion of individuals with LBP initially seek treatment from primary care providers (PCP) and physician specialists with management emphasizing second- and third-line approaches.^3^ Non-prescribing HCPs, like chiropractors (DC), physical therapists (PT), or licensed acupuncturists (LAc) are more likely to have episodes associated with an emphasis on first-line therapies, with less use of second- and third-line services.^3,6^ A variety of confounders impact the interpretation and translation of findings associated with the type of HCP initially contacted by an individual with LBP.^4,6,7^

Relatively little is known about longitudinal patterns of all types of HCPs involved in an episode of LBP and association with guideline concordance and total cost. Research into the sequence of types of HCPs involved in an episode of LBP has tended to focus on service use rather than types of HCPs involved, and has often focused on a single type of initial contact HCP such as PCP ^9,10^, physiatry ^11^, emergency medicine (EM)^12,13^, and PT.^5,8,14^ These studies identified variation in approaches to LBP, yet have not provide comprehensive insights into patterns of HCP involvement in LBP. A PubMed.gov search for key terms “last provider seen” and “back pain” returned no results.

The aim of this study was to examine patterns in the types of HCP involved in a single episode of non-surgical LBP, exposure to 13 types of health care services, and total cost, with a specific focus on association with the type of HCP seen last. The hypothesis was types of HCP emphasizing CPG concordant first-line approaches, like DCs, PTs, LAcs, and doctors of osteopathy (DO), would be commonly seen as the last HCP during an episode of LBP even if becoming involved later in an episode.

## Methods

### Study design, population, setting and data sources

This is a retrospective cohort study of individuals with a single episode of non-surgical LBP during the 2017-2019 study period. An analytic database was created which included de-identified enrollment records, administrative claims data for all inpatient and outpatient services, and pharmacy prescriptions, for commercially insured enrollees from a single national commercial insurer. De-identified HCP demographic information and professional licensure status was included in an HCP database. ZIP code level population race and ethnicity data was extracted from the US Census Bureau ^15^, socioeconomic status (SES) Area Deprivation Index (ADI) data, from the University of Wisconsin Neighborhood Atlas^® 16^, and household adjusted gross income (AGI) from the Internal Revenue Service.^17^

The study data was de-identified or a Limited Data Set in compliance with the Health Insurance Portability and Accountability Act and customer requirements. The UnitedHealth Group Office of Human Research Affairs determined that this study was exempt from Institutional Review Board review. The study was conducted and reported based on the Strengthening the Reporting of Observational Studies in Epidemiology (STROBE) guidelines.^18^ [Supplement 1 – STROBE Checklist]

The study does not include an adjustment for typical confounders such as individual age, sex and co-morbidities ^19,20^ using common yet potentially inadequate approaches such as propensity score matching. ^21^ The first reason for this approach is an inability to control for all measurable and unmeasured confounders, and confounders of known confounders, of the selection of the initial and subsequent HCPs involved in an episode of LBP. Examples of confounders not available include: HCP options convenient to an individual’s home, workplace or daily travel routes including public transportation if used, individual preference for specific services or type of HCP including gender or racial concordance, recommendations from family or friends and influence of HCP marketing efforts, perceived LBP severity, anticipated potential out of pocket costs and willingness to pay for services, time availability to participate in multiple visits, and appointment availability within an individual’s timing expectations meeting these and other criteria.^22^ The second reason is to avoid blurring the line between association and causality through a process that simultaneously introduces distortion and complexity into results. The study aim is to produce actual measures of individual demographic and episodic characteristics, and unadjusted associations, that are reported for each type of HCP seen initially, or subsequently, during an episode of LBP.

As an alternative to incomplete confounding control, the cohort was limited to individuals with a single episode of non-surgical LBP, and sub-analyses were performed for episodes where an individual initially contacted a PCP, orthopedic surgeon (OS), or EM physician. To examine the influence of population factors on results, the flow of episodes between types of HCP, and the of % of episodes where each type of HCP was the last HCP seen were calculated for individuals living in zip codes with 75-100 ADI and 0-25% non-Hispanic white (NHW) population, 50-75 ADI and 50-75% NHW population, and 0-25 ADI and 75-100% NWH population.

### Cohort selection and unit of analysis

The cohort included individuals aged 18 years and older with a single complete episode of non-surgical LBP commencing and ending during the calendar years 2017-2019. This timeframe was selected to follow the release of the American College of Physicians (ACP) LBP CPG ^2^ in 2017 and before the influence of the COVID-19 epidemic on care patterns in early 2020. All individuals had continuous medical and pharmacy commercial insurance coverage during the entire study period.

Episode of care was selected as the unit of analysis. This approach has been shown to be a valid way to organize all administrative claims data associated with a condition.^23^ The *Symmetry*^*®*^ *Episode Treatment Groups*^*®*^ *(ETG*^*®*^) and *Episode Risk Groups*^*®*^ *(ERG*^*®*^) version 9.5 methodologies and definitions were used to translate administrative claims data into episodes of care, which have been reported as a valid measurement for comparison of HCPs based on cost of care.^24^ A previous analysis of the same underlying data indicates a low risk of misclassification bias associated with using episode of care as the unit of analysis.^3^ Using episode of care unit of measurement has potential translational benefits in supporting the transition from fee for service to value-based episodic bundled payment arrangements, something particularly relevant when examining episodes with multiple HCPs involved.

The analysis included complete episodes defined as having at least 91-day pre- and 61-day post-episode clean periods during which no services were provided by any HCP for any LBP diagnosis. LBP episodes including a surgical procedure, or associated with diagnoses of malignant and non-malignant neoplasms, fractures and other spinal trauma, infection, congenital deformities and scoliosis, autoimmune disorders, osteoporosis, and advanced arthritis were excluded from the analysis. Individuals with multiple LBP episodes during the study period were also excluded. These exclusions were made to address a potential study limitation of individuals with more complex conditions confounding study results examining the type of HCP initially contacted, and subsequent types of HCP seen during an episode of LBP.

### Variables

Data preprocessing, table generation, and initial analyses were performed using Python (*Python Language Reference, Version 3*.*7*.*5*., n.d.). To evaluate whether measures were derived from a normally distributed sample we conducted a goodness-of-fit measure using D’Agostino’s K-squared test. Non-normally distributed data are reported using the median and interquartile range (IQR). Odds (OR) and risk (RR) ratios, and associated 95% confidence intervals (CI), were calculated for select measures. RR were reported as this is the measure more widely understood in associational analyses and due to the tendency for ORs to exaggerate risk in situations where an outcome is relatively common.^25^

The primary independent variable was 17 types of HCP initially contacted by an individual experiencing a single episode of non-surgical LBP during the study period. All types of HCP could be accessed directly without a referral. The primary dependent variables were types of HCP involved after the initial HCP, the number of days into an episode when subsequent HCP types were involved, the rate of use of 13 types of health care services and total cost. Service utilization reflects services provided by any type of HCP an individual saw during the complete episode of LBP. Total cost included costs associated with all services provided for LBP during an episode, including those not specifically identified in the 13 categories used in the analyses. Costs for services for which an insurance claim was not submitted were not available.

With 97.5% of episodes involving five or fewer HCPs, the analyses focused on the first five types of HCP seen during an episode. This resulted in 1,419,925 possible HCP sequence combinations. [Figure 1] Rate (%) and timing (days), median and IQR, and RR and 95% CI results for cohort demographics, HCP involvement, episodic service use, and total cost were calculated for all HCP sequence combinations. Total episode cost and percent of episodes including an MRI or prescription opioid were calculated for episodes where a PCP, DC, PT or OS was the last HCP seen.

**Figure 1.**
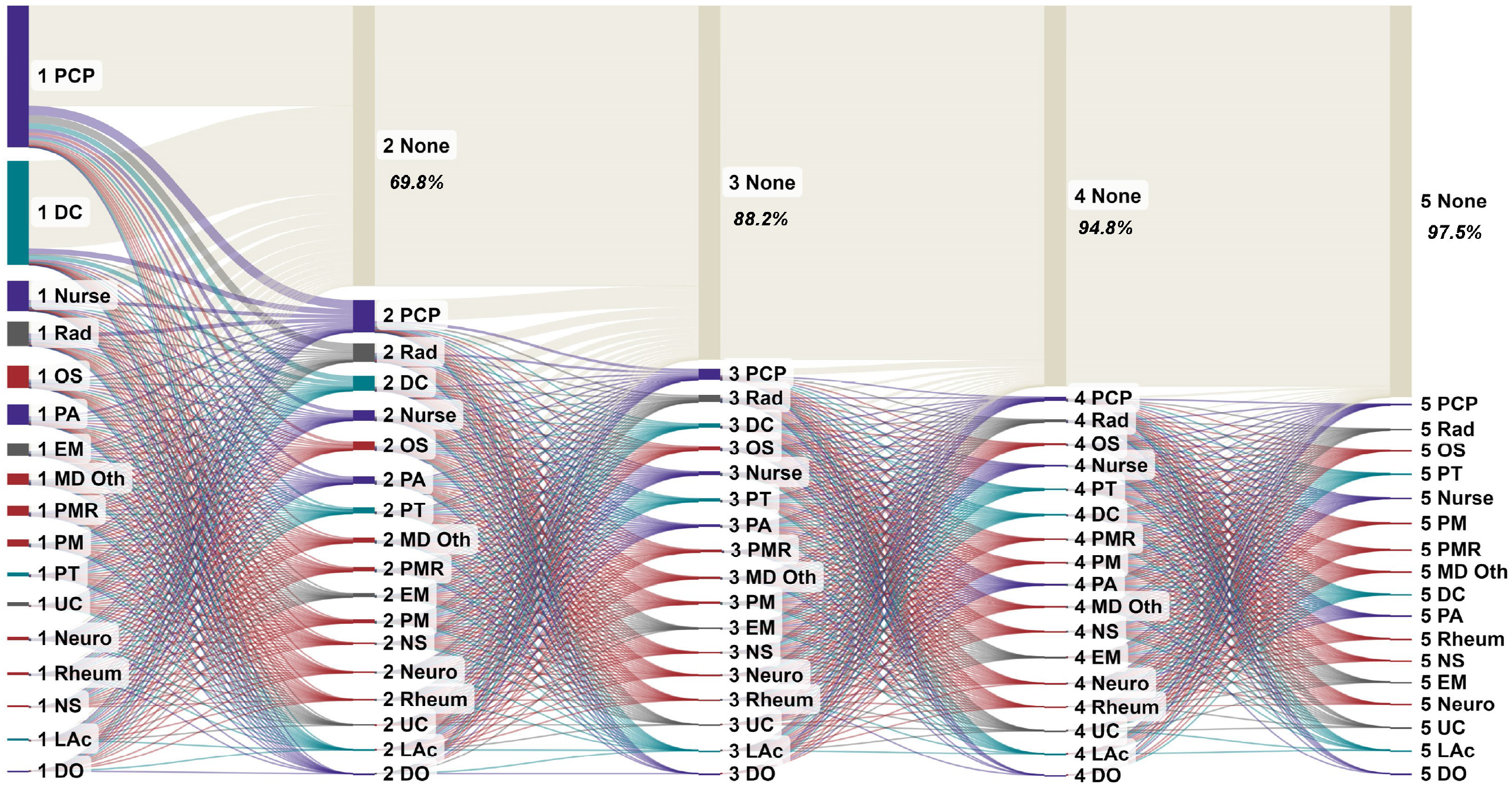
Flow of episodes from initial (1) to second (2) to third (3) to fourth (4) to fifth (5) type of health care provider (HCP) seen during an episode PCP = primary care provider, DC=doctor of chiropractic, Rad=radiologist, OS=orthopedic surgeon, PA=physician’s assistant, EM=emergency medicine, oth=other, PMR=physical medicine & rehabilitation, PM=pain management, PT=physical therapist, UC=urgent care Neuro=neurologist, Rheum=rheumatologist, NS=neurosurgeon, LAc=licensed acupuncturist, DO=doctor of osteopathy, None = no next HCP seen (previous HCP was the last HCP)

## Results

The cohort consisted of 503,958 individuals, associated with 503,958 complete non-surgical LBP episodes for which 196,522 different HCPs were initially contacted. There were $387,867,096 in reimbursed health care expenditures. The median pre- (646 days) and post-episode (405 days) clean periods were substantially longer than the *ETG*^*®*^ clean period definitions. Primary care physicians (PCP) (35.2% of episodes), DCs (25.9%), nurse practitioners (7.5%), radiologists (6.0%) and OSs (5.5%) where the most common types of HCP initially contacted. [Supplement 1] [Supplement 2] There were potentially meaningful differences in individual and population factors between types of HCP seen at different points in an episode. [Supplement 3]

30.2% of episodes involved 2 HCPs, 11.8% involved 3 HCPs, 5.2% involved 4 HCPs and 2.5% of episodes involved 5 HCPs. All types of HCP were involved in progressively fewer episodes when moving from initial to second, third, fourth or fifth HCP, except PTs who participated in more episodes as a second HCP (7,344) than as an initial HCP (5,114). [Table 1]

**Table 1.** Non-surgical low back pain episode count by type of health care provider (HCP) and episode sequence position

| HCP type |  | # of Unique HCPs |  |  |  |  | # of Episodes |  |  |  |  | % of Episodes |  |  |  |  |
| --- | --- | --- | --- | --- | --- | --- | --- | --- | --- | --- | --- | --- | --- | --- | --- | --- |
|  |  | First HCP | Second HCP | Third HCP | Fourth HCP | Fifth HCP | First HCP | Second HCP | Third HCP | Fourth HCP | Fifth HCP | First HCP | Second HCP | Third HCP | Fourth HCP | Fifth HCP |
| Primary Care | PCP | 68888 | 28730 | 11787 | 4889 | 2301 | 177265 | 40767 | 13766 | 5239 | 2364 | 35.2% | 26.8% | 23.2% | 20.0% | 18.5% |
|  | Nurse | 21226 | 9897 | 4101 | 1919 | 1049 | 37857 | 12820 | 4749 | 2108 | 1031 | 7.5% | 8.4% | 8.0% | 8.0% | 8.1% |
|  | PA | 12787 | 6746 | 2955 | 1373 | 781 | 25350 | 9418 | 3542 | 1545 | 718 | 5.0% | 6.2% | 6.0% | 5.9% | 5.6% |
|  | DO | 121 | 45 | 30 | 9 | 7 | 469 | 64 | 42 | 11 | 10 | 0.1% | 0.0% | 0.1% | 0.0% | 0.1% |
| Non-Prescriber | DC | 21644 | 9953 | 4396 | 1735 | 739 | 130520 | 18586 | 5823 | 2011 | 797 | 25.9% | 12.2% | 9.8% | 7.7% | 6.2% |
|  | PT | 3019 | 4143 | 2769 | 1601 | 776 | 5114 | 7344 | 4076 | 2077 | 1135 | 1.0% | 4.8% | 6.9% | 7.9% | 8.9% |
|  | LAc | 1430 | 397 | 159 | 67 | 36 | 2634 | 486 | 175 | 73 | 30 | 0.5% | 0.3% | 0.3% | 0.3% | 0.2% |
| Specialist | OS | 8590 | 5241 | 3048 | 1740 | 1120 | 27799 | 10867 | 4971 | 2362 | 1152 | 5.5% | 7.1% | 8.4% | 9.0% | 9.0% |
|  | PMR | 2966 | 2127 | 1622 | 1120 | 767 | 12496 | 5501 | 3236 | 1817 | 1009 | 2.5% | 3.6% | 5.5% | 6.9% | 7.9% |
|  | PM | 2497 | 1888 | 1561 | 1126 | 782 | 9013 | 4280 | 2884 | 1757 | 1031 | 1.8% | 2.8% | 4.9% | 6.7% | 8.1% |
|  | NS | 1373 | 1164 | 894 | 650 | 441 | 3058 | 2084 | 1322 | 817 | 432 | 0.6% | 1.4% | 2.2% | 3.1% | 3.4% |
|  | Neuro | 2352 | 1321 | 709 | 371 | 194 | 3917 | 1729 | 843 | 410 | 224 | 0.8% | 1.1% | 1.4% | 1.6% | 1.8% |
|  | Rheum | 1642 | 998 | 558 | 304 | 158 | 3354 | 1463 | 667 | 332 | 166 | 0.7% | 1.0% | 1.1% | 1.3% | 1.3% |
|  | MD Oth | 9230 | 5122 | 2332 | 1259 | 723 | 13877 | 6716 | 2963 | 1539 | 840 | 2.8% | 4.4% | 5.0% | 5.9% | 6.6% |
| Emergency /Urgent Care | EM | 8365 | 3658 | 1313 | 511 | 301 | 15760 | 4838 | 1463 | 544 | 233 | 3.1% | 3.2% | 2.5% | 2.1% | 1.8% |
|  | Rad | 10098 | 8613 | 4763 | 2468 | 2039 | 30393 | 23828 | 8454 | 3475 | 1551 | 6.0% | 15.7% | 14.3% | 13.3% | 12.2% |
|  | UC | 1120 | 517 | 186 | 73 | 43 | 5082 | 1358 | 272 | 87 | 39 | 1.0% | 0.9% | 0.5% | 0.3% | 0.3% |
| Total |  | 177348 | 90560 | 43183 | 21215 | 12257 | 503958 | 152149 | 59248 | 26204 | 12762 | 100.0% | 100.0% | 100.0% | 100.0% | 100.0% |
| % of Total Episodes |  |  |  |  |  |  |  |  |  |  |  | 100.0% | 30.2% | 11.8% | 5.2% | 2.5% |

PCP=primary care provider, DO=doctor of osteopathy, DC=doctor of chiropractic, PT=physical therapist, LAc=licensed acupuncturist, OS=orthopedic surgeon, PMR=physical medicine & rehabilitation, PM=pain medicine, NS=neurosurgeon, Neuro=neurologist, Rheum=rheumatologist, MD Oth = other medical physician, EM=emergency medicine, Rad=radiologist, UC=urgent care

When initially contacted by individuals with LBP, and compared to the PCP reference (71.0% of episodes), DCs (84.0%, risk ratio 1.18, 95% confidence interval 1.18-1.19), LAcs (81.7%, 1.15, 1.13-1.17) and DOs (80.2%, 1.13, 1.08, 1.18) were most likely to be the last and only HCP seen during an episode. [Table 2] [Figure 2] When seen as a second, third or fourth HCP during an episode, DCs and PTs were the only HCPs more likely than a PCP to be the last HCP seen. Neurosurgeons and radiologists were the least likely to be the last HCP seen. Sub analyses of the 75-100 ADI 0-25% NHW, 50-75 ADI 50-75% NHW, and 0-25 ADI 75-100% NHW segments revealed nearly identical results. [Supplement 4] When the initial HCP contacted, DCs were more likely than PCPs to be the last HCP seen in each ADI NHW segment. LAcs were more likely than PCPs to be the last HCP seen in the 75-100 ADI 0-25% NHW and 0-25 ADI 75-100% NHW segments. [Figure 3] When positioned as the second HCP involved in an episode, DCs were more likely than PCPs to be the last HCP seen in each segment. PTs were more likely than PCPs to be the last HCP seen in the 50-75 ADI 50-75% NHW and 0-25 ADI 75-100% NHW segments. [Figure 4]

**Table 2.**
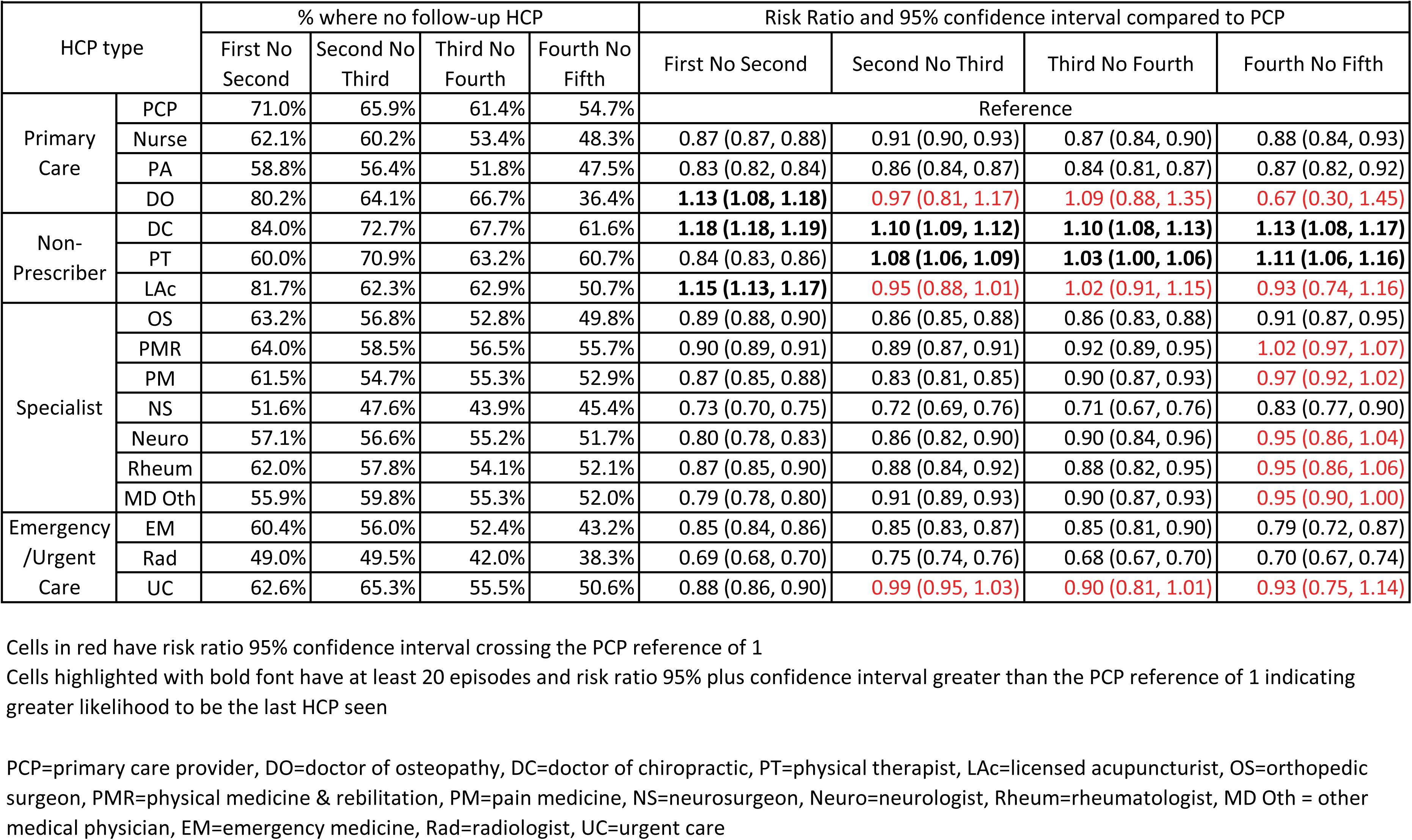
% of non-surgical low back pain episodes where health care provider (HCP) type was the last HCP seen

| HCP type |  | % where no follow-up HCP |  |  |  | Risk Ratio and 95% confidence interval compared to PCP |  |  |  |
| --- | --- | --- | --- | --- | --- | --- | --- | --- | --- |
|  |  | First No Second | Second No Third | Third No Fourth | Fourth No Fifth | First No Second | Second No Third | Third No Fourth | Fourth No Fifth |
| Primary Care | PCP | 71.0% | 65.9% | 61.4% | 54.7% | Reference |  |  |  |
|  | Nurse | 62.1% | 60.2% | 53.4% | 48.3% | 0.87 (0.87, 0.88) | 0.91 (0.90, 0.93) | 0.87 (0.84, 0.90) | 0.88 (0.84, 0.93) |
|  | PA | 58.8% | 56.4% | 51.8% | 47.5% | 0.83 (0.82, 0.84) | 0.86 (0.84, 0.87) | 0.84 (0.81, 0.87) | 0.87 (0.82, 0.92) |
|  | DO | 80.2% | 64.1% | 66.7% | 36.4% | **1.13 (1.08, 1.18)** | **0.97 (0.81, 1.17)** | **1.09 (0.88, 1.35)** | **0.67 (0.30, 1.45)** |
| Non-Prescriber | DC | 84.0% | 72.7% | 67.7% | 61.6% | **1.18 (1.18, 1.19)** | **1.10 (1.09, 1.12)** | **1.10 (1.08, 1.13)** | **1.13 (1.08, 1.17)** |
|  | PT | 60.0% | 70.9% | 63.2% | 60.7% | 0.84 (0.83, 0.86) | **1.08 (1.06, 1.09)** | **1.03 (1.00, 1.06)** | **1.11 (1.06, 1.16)** |
|  | LAc | 81.7% | 62.3% | 62.9% | 50.7% | **1.15 (1.13, 1.17)** | **0.95 (0.88, 1.01)** | **1.02 (0.91, 1.15)** | **0.93 (0.74, 1.16)** |
| Specialist | OS | 63.2% | 56.8% | 52.8% | 49.8% | 0.89 (0.88, 0.90) | 0.86 (0.85, 0.88) | 0.86 (0.83, 0.88) | 0.91 (0.87, 0.95) |
|  | PMR | 64.0% | 58.5% | 56.5% | 55.7% | 0.90 (0.89, 0.91) | 0.89 (0.87, 0.91) | 0.92 (0.89, 0.95) | **1.02 (0.97, 1.07)** |
|  | PM | 61.5% | 54.7% | 55.3% | 52.9% | 0.87 (0.85, 0.88) | 0.83 (0.81, 0.85) | 0.90 (0.87, 0.93) | **0.97 (0.92, 1.02)** |
|  | NS | 51.6% | 47.6% | 43.9% | 45.4% | 0.73 (0.70, 0.75) | 0.72 (0.69, 0.76) | 0.71 (0.67, 0.76) | 0.83 (0.77, 0.90) |
|  | Neuro | 57.1% | 56.6% | 55.2% | 51.7% | 0.80 (0.78, 0.83) | 0.86 (0.82, 0.90) | 0.90 (0.84, 0.96) | **0.95 (0.86, 1.04)** |
|  | Rheum | 62.0% | 57.8% | 54.1% | 52.1% | 0.87 (0.85, 0.90) | 0.88 (0.84, 0.92) | 0.88 (0.82, 0.95) | **0.95 (0.86, 1.06)** |
|  | MD Oth | 55.9% | 59.8% | 55.3% | 52.0% | 0.79 (0.78, 0.80) | 0.91 (0.89, 0.93) | 0.90 (0.87, 0.93) | **0.95 (0.90, 1.00)** |
| Emergency /Urgent Care | EM | 60.4% | 56.0% | 52.4% | 43.2% | 0.85 (0.84, 0.86) | 0.85 (0.83, 0.87) | 0.85 (0.81, 0.90) | 0.79 (0.72, 0.87) |
|  | Rad | 49.0% | 49.5% | 42.0% | 38.3% | 0.69 (0.68, 0.70) | 0.75 (0.74, 0.76) | 0.68 (0.67, 0.70) | 0.70 (0.67, 0.74) |
|  | UC | 62.6% | 65.3% | 55.5% | 50.6% | 0.88 (0.86, 0.90) | **0.99 (0.95, 1.03)** | **0.90 (0.81, 1.01)** | **0.93 (0.75, 1.14)** |

Cells in red have risk ratio 95% confidence interval crossing the PCP reference of 1
Cells highlighted with bold font have at least 20 episodes and risk ratio 95% plus confidence interval greater than the PCP reference of 1 indicating greater likelihood to be the last HCP seen
PCP=primary care provider, DO=doctor of osteopathy, DC=doctor of chiropractic, PT=physical therapist, LAc=licensed acupuncturist, OS=orthopedic surgeon, PMR=physical medicine & rehabilitation, PM=pain medicine, NS=neurosurgeon, Neuro=neurologist, Rheum=rheumatologist, MD Oth = other medical physician, EM=emergency medicine, Rad=radiologist, UC=urgent care

**Figure 2.**
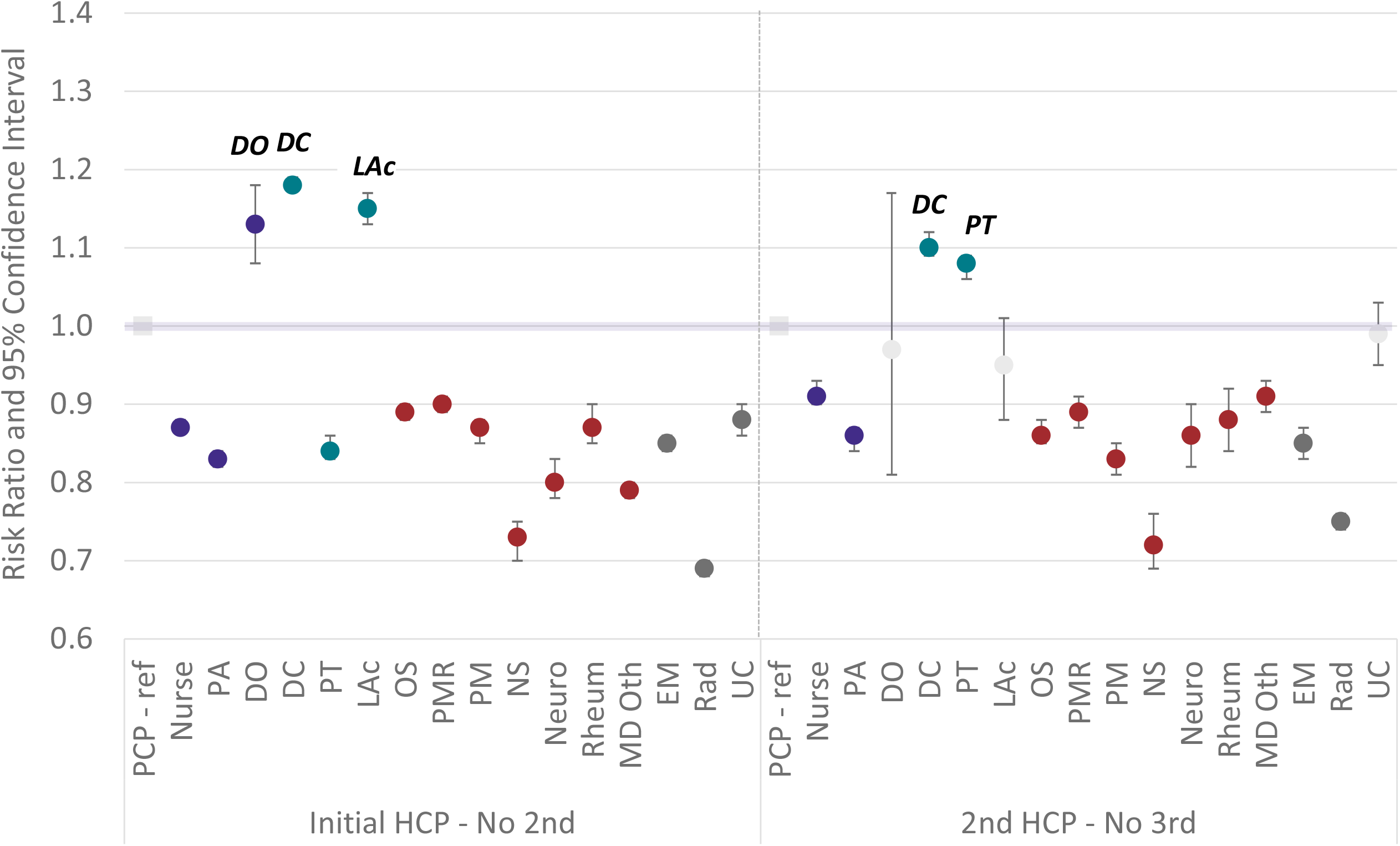
Risk ratio and 95% confidence interval compared to primary care provider (PCP) reference for % of episodes where HCPs with at least 20 episodes were the initial and last or 2nd and last HCP. PA=physician’s assistant, DO=doctor of osteopathy, DC=doctor of chiropractic, PT=physical therapist, LAc=licensed acupuncturist, OS=orthopedic surgeon, PMR=physical medicine & rehabilitation, PM=pain management, NS=neurosurgeon, Neuro=neurologist, Rheum=rheumatologist, Oth=other, EM=emergency medicine, Rad=radiologist, UC=urgent care

**Figure 3.**
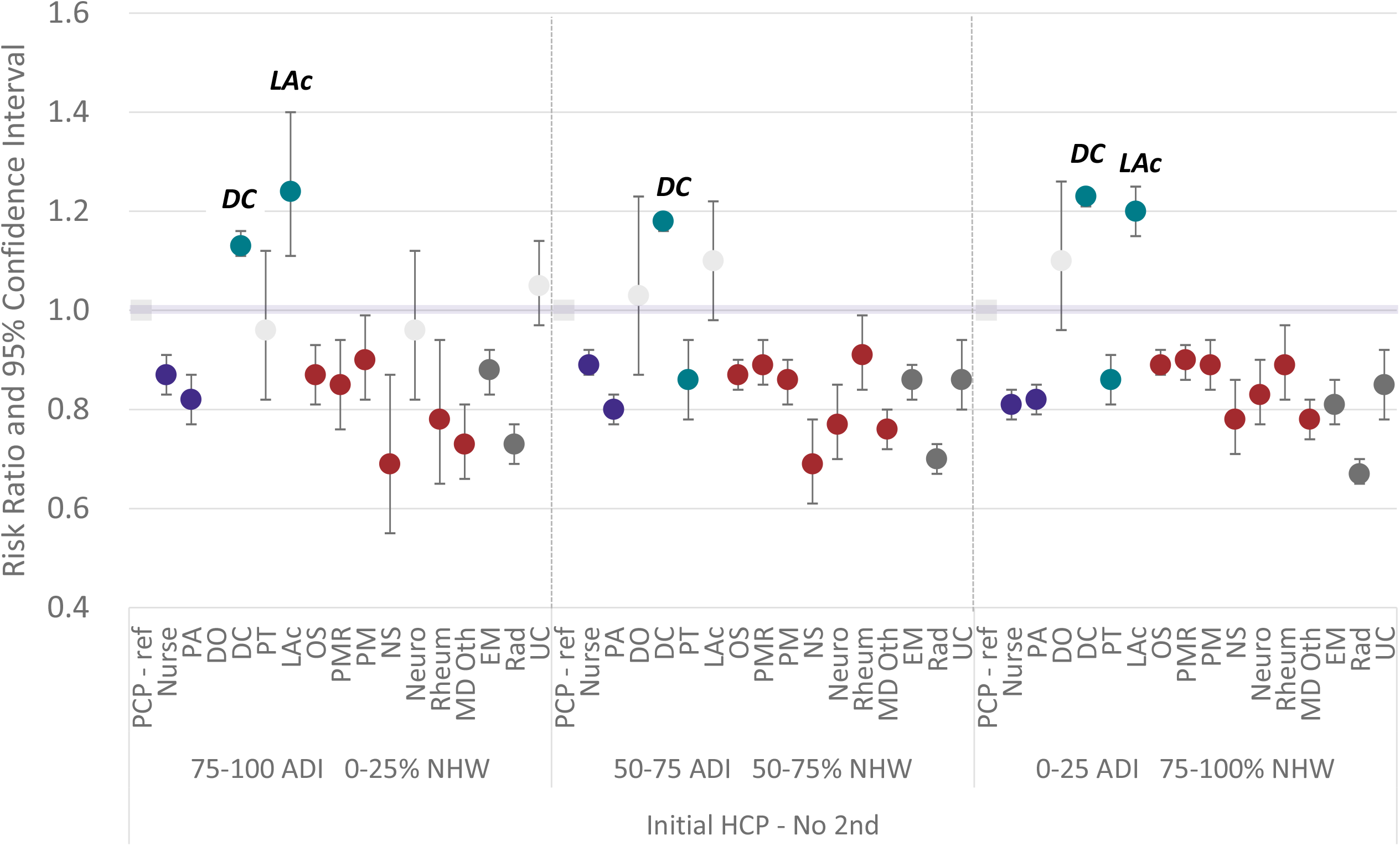
Risk ratio and 95% confidence interval compared to primary care provider (PCP) reference for % of episodes where HCPs with at least 20 episodes were the initial and last HCP seen segmented by population Area Deprivation Index (ADI) and % of the population that is non-Hispanic white (NHW). PA=physician’s assistant, DO=doctor of osteopathy, DC=doctor of chiropractic, PT=physical therapist, LAc=licensed acupuncturist, OS=orthopedic surgeon, PMR=physical medicine & rehabilitation, PM=pain management, NS=neurosurgeon, Neuro=neurologist, Rheum=rheumatologist, Oth=other, EM=emergency medicine, Rad=radiologist, UC=urgent care

**Figure 4.**
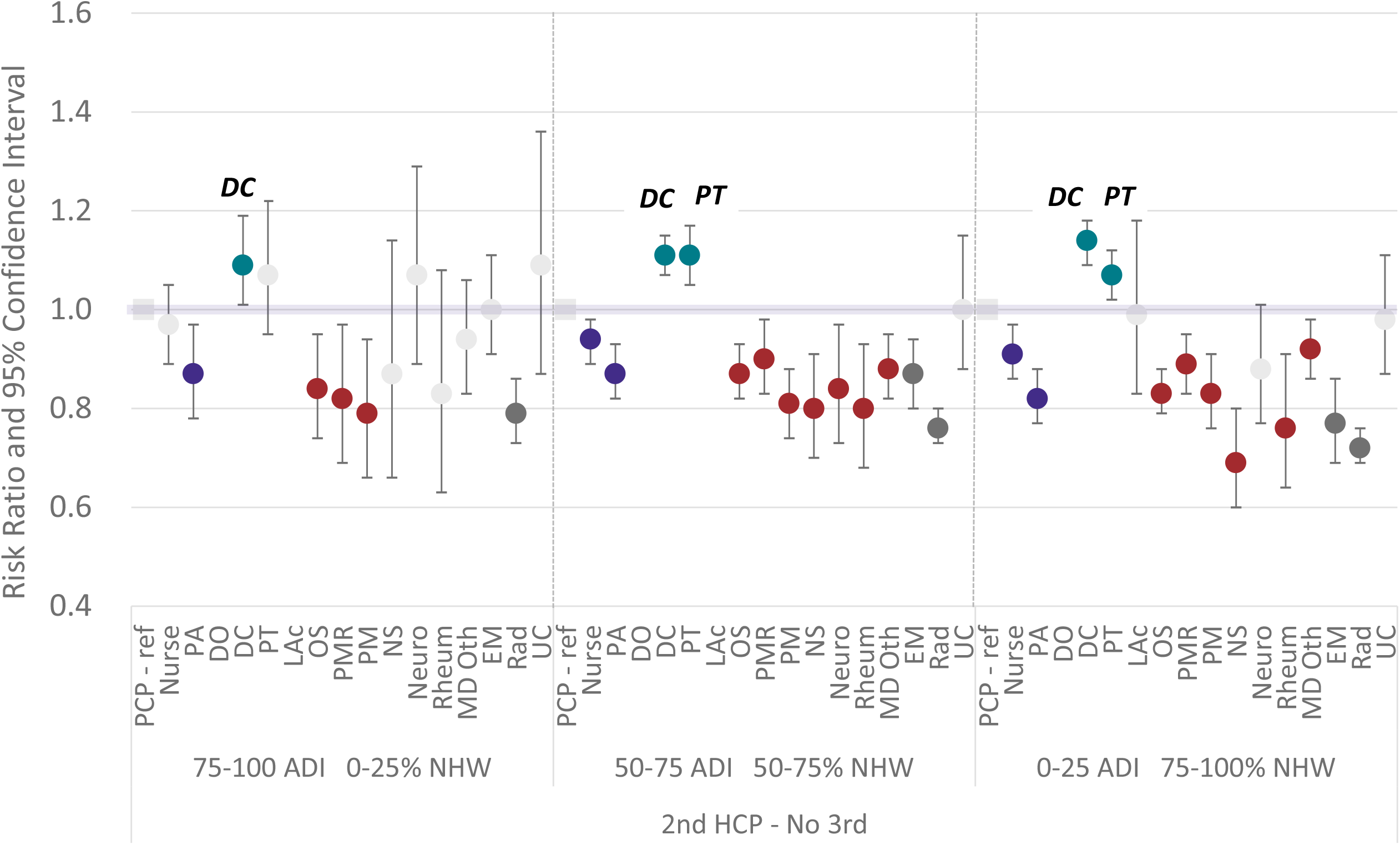
Risk ratio and 95% confidence interval compared to primary care provider (PCP) reference for % of episodes where HCPs with at least 20 episodes were the second and last HCP seen segmented by population Area Deprivation Index (ADI) and % of the population that is non-Hispanic white (NHW). PA=physician’s assistant, DO=doctor of osteopathy, DC=doctor of chiropractic, PT=physical therapist, LAc=licensed acupuncturist, OS=orthopedic surgeon, PMR=physical medicine & rehabilitation, PM=pain management, NS=neurosurgeon, Neuro=neurologist, Rheum=rheumatologist, Oth=other, EM=emergency medicine, Rad=radiologist, UC=urgent care

The sub-analyses of individuals initially contacting a PCP, OS, or EM had similar last HCP seen results. For individuals initially contacting a PCP, and when seen as a second HCP, DCs (76.2%, risk ratio 1.15, 95% confidence interval 1.13-1.18) and PTs (76.1%, 1.15, 1.12-1.18) were the most likely to be the last HCP seen. For individuals initially contacting an OS, and when seen as a second HCP, DCs (69.5%, 1.12, 1.05-1.19) and PTs (71.2%, 1.14, 1.09-1.20) were the most likely to be the last HCP seen. For individuals initially contacting EM, and when seen as a second HCP DCs (76.5%, 1.12, 1.05-1.20) were the most likely to be the last HCP seen. [Figure 5] This was also the case when a DC or PT was the third or fourth provider seen for individuals initially contacting a PCP. DCs and PTs were also the most likely to be the last HCP seen when the second, third or fourth HCP for individuals initially contacting an OS or EM. [Table 3]

**Table 3.**
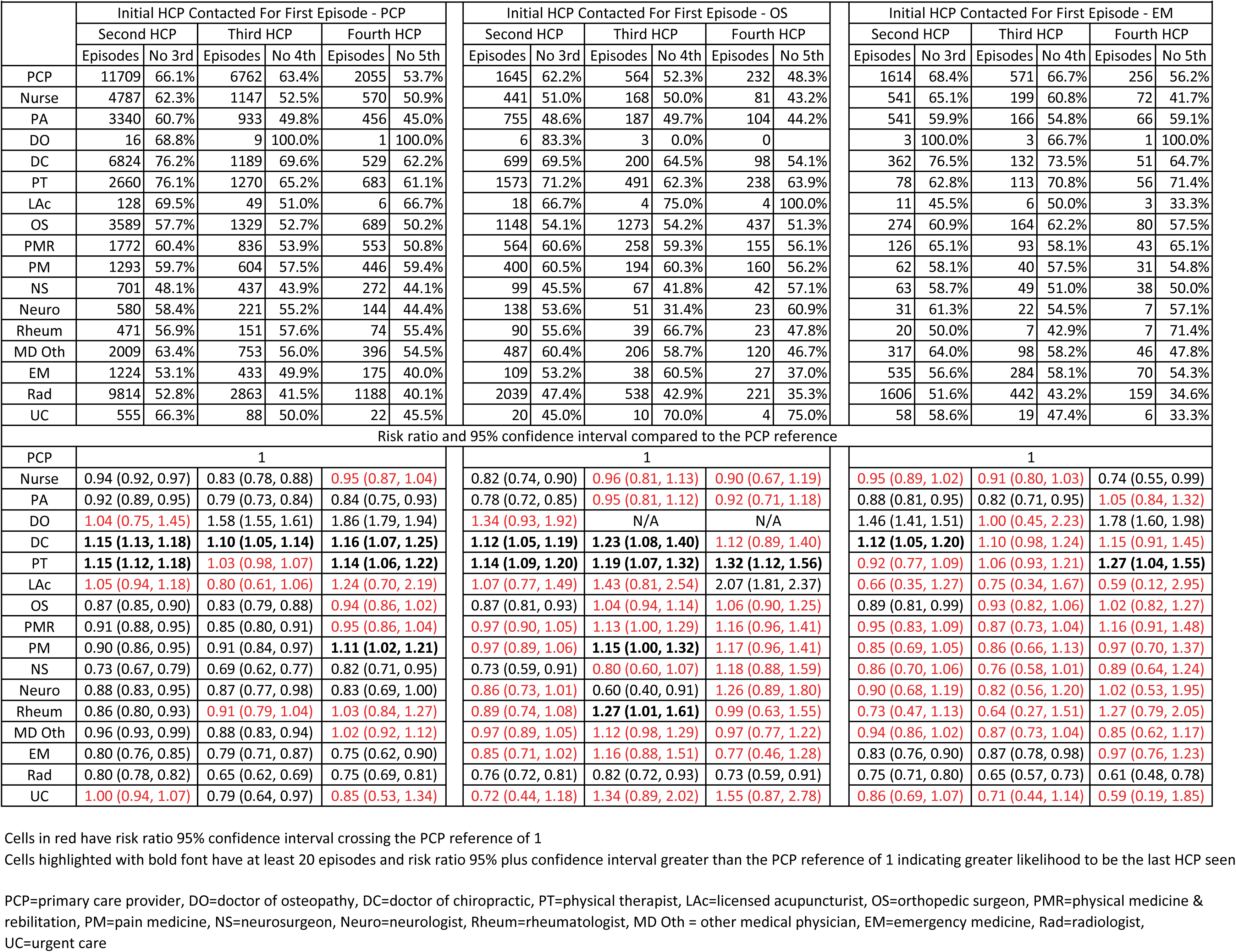
Non-surgical low back pain episodes initially contacting a primary care provider (PCP), orthopedic surgeon (OS) or emergency medicine physician (EM), subsequent health care providers (HCP) seen, and % of episodes where the subsequent HCP was the last HCP seen

**Figure 5.**
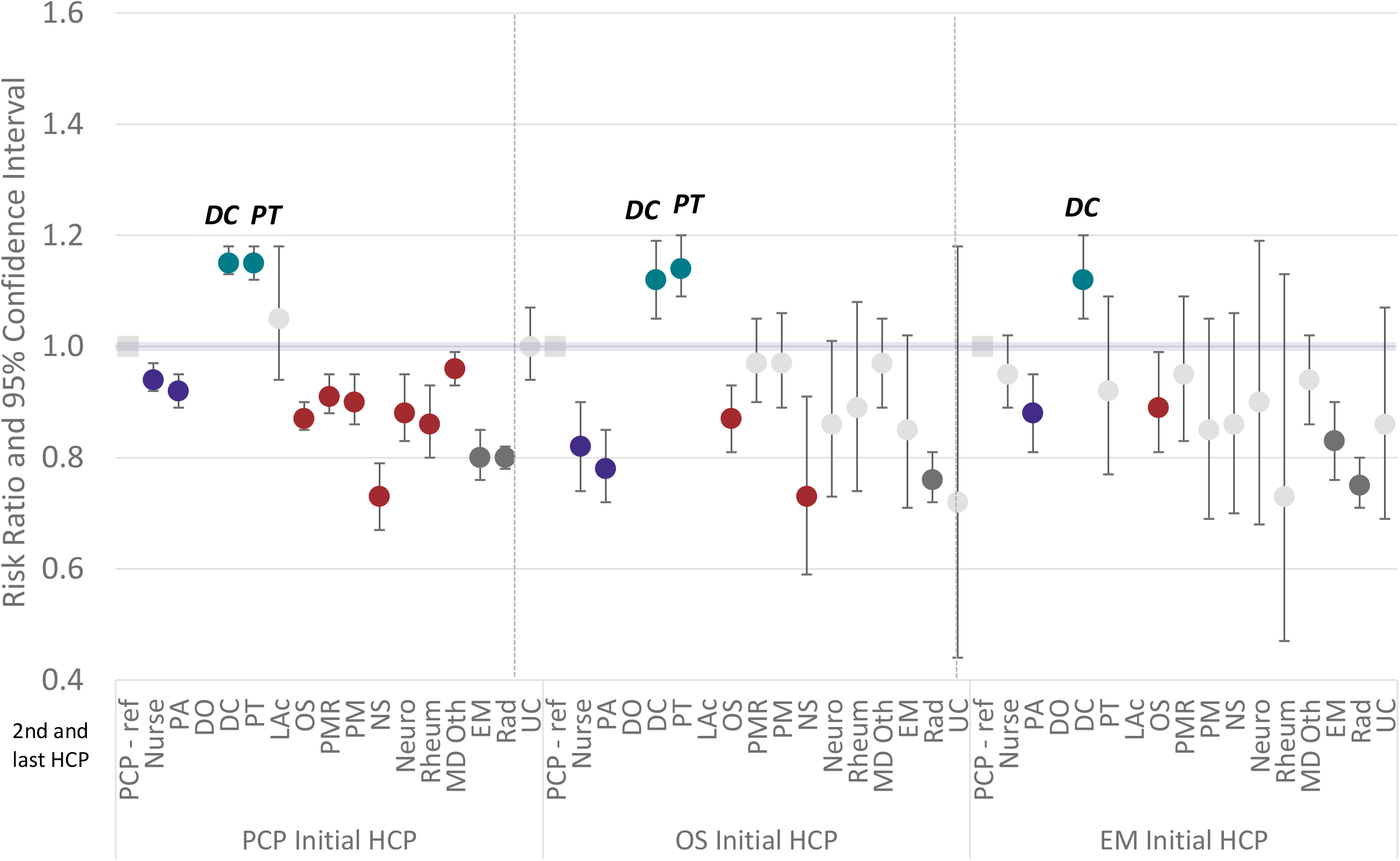
- Risk ratio and 95% confidence interval compared to primary care provider (PCP) reference for % of episodes where for health care providers with at least 20 episodes the type of second HCP seen was also the last HCP seen. PA=physician’s assistant, DO=doctor of osteopathy, DC=doctor of chiropractic, PT=physical therapist, LAc=licensed acupuncturist, OS=orthopedic surgeon, PMR=physical medicine & rehabilitation, PM=pain management, NS=neurosurgeon, Neuro=neurologist, Rheum=rheumatologist, Oth=other, EM=emergency medicine, Rad=radiologist, UC=urgent care

When the last HCP seen during an episode the rate of MRI and opioid exposure progressively increased the later in an episode a PCP, DC, PT or OS became involved. Compared to both PCPs and OSs, DCs and PTs were associated with a lower rate of opioid exposure when becoming involved at any time during an episode. DCs were associated with a lower rate of MRI exposure than PCPs, PTs, or OSs. [Figure 6] Similarly, when the last HCP seen, total episode cost progressively increased the later in an episode a PCP, DC, PT or OS became involved. When the last HCP seen, PCPs and DCs were associated with lower median total episode cost with PTs and OSs associated with higher median total episode cost. [Figure 7] [Table 4]

**Table 4.** Non-surgical low back pain service use and total cost for select health care providers (HCP) positioned as the last HCP seen

|  | Primary care provider (PCP) |  |  |  | Doctor of Chiropractic (DC) |  |  |  | Physical Therapist (PT) |  |  |  | Orthopedic Surgeon (OS) |  |  |  |
| --- | --- | --- | --- | --- | --- | --- | --- | --- | --- | --- | --- | --- | --- | --- | --- | --- |
| Episode Sequence | 1st | 2nd | 3rd | 4th | 1st | 2nd | 3rd | 4th | 1st | 2nd | 3rd | 4th | 1st | 2nd | 3rd | 4th |
|  | No 2nd | No 3rd | No 4th | No 5th | No 2nd | No 3rd | No 4th | No 5th | No 2nd | No 3rd | No 4th | No 5th | No 2nd | No 3rd | No 4th | No 5th |
| Episodes | 88980 | 24766 | 8251 | 2835 | 107340 | 13409 | 3928 | 1235 | 3030 | 5177 | 2572 | 1261 | 15314 | 5965 | 2593 | 1172 |
| % of episodes including service |  |  |  |  |  |  |  |  |  |  |  |  |  |  |  |  |
| Manipulation- Chiropractic | 4.1% | 20.7% | 18.4% | 20.9% | 93.6% | 91.6% | 94.8% | 93.9% | 7.8% | 6.6% | 10.1% | 12.5% | 3.1% | 11.6% | 13.5% | 14.9% |
| Active Care | 9.8% | 15.1% | 21.2% | 28.3% | 30.9% | 35.5% | 43.4% | 47.9% | 96.6% | 97.2% | 97.3% | 97.9% | 20.1% | 27.6% | 35.2% | 42.2% |
| Passive Therapy | 4.5% | 10.3% | 11.9% | 14.8% | 36.8% | 38.9% | 46.3% | 50.3% | 28.7% | 26.5% | 28.5% | 30.3% | 6.7% | 11.0% | 14.3% | 14.2% |
| Manual Therapy | 7.3% | 10.3% | 14.0% | 18.8% | 17.9% | 21.0% | 27.3% | 33.2% | 77.4% | 74.2% | 73.8% | 74.8% | 14.1% | 18.8% | 24.0% | 28.5% |
| Acupuncture | 0.3% | 0.6% | 0.7% | 0.7% | 0.5% | 1.2% | 1.9% | 2.3% | 1.2% | 0.6% | 0.9% | 1.0% | 0.2% | 0.5% | 0.6% | 0.7% |
| Manipulation - Osteopathic | 2.5% | 1.3% | 1.6% | 1.5% | 0.1% | 0.4% | 0.5% | 0.4% | 0.3% | 0.4% | 0.7% | 0.6% | 0.7% | 0.4% | 0.8% | 0.5% |
| Imaging - Radiography | 21.1% | 31.6% | 45.7% | 51.5% | 18.8% | 29.5% | 43.0% | 53.6% | 16.3% | 42.7% | 57.7% | 63.4% | 73.6% | 68.7% | 71.9% | 73.5% |
| Rx - NSAID | 44.2% | 42.7% | 49.5% | 52.6% | 4.0% | 20.1% | 24.0% | 30.9% | 9.9% | 22.4% | 30.8% | 33.0% | 25.8% | 39.0% | 44.2% | 50.1% |
| Rx - Skeletal Muscle Relaxant | 47.5% | 43.3% | 46.7% | 47.9% | 2.3% | 16.1% | 22.3% | 27.0% | 9.3% | 22.8% | 29.2% | 32.8% | 15.8% | 26.6% | 33.2% | 36.3% |
| Imaging - MRI | 4.9% | 6.5% | 13.3% | 24.8% | 1.2% | 3.7% | 8.8% | 16.8% | 9.6% | 11.4% | 22.2% | 33.0% | 18.5% | 27.8% | 40.2% | 50.5% |
| Rx-Opioid | 26.4% | 26.9% | 35.9% | 39.9% | 3.4% | 11.4% | 15.6% | 21.1% | 6.6% | 8.6% | 13.7% | 19.2% | 12.5% | 20.0% | 28.1% | 33.4% |
| Spinal Injection | 2.5% | 2.7% | 4.6% | 7.9% | 0.6% | 2.0% | 3.8% | 5.4% | 3.4% | 3.3% | 5.4% | 10.2% | 7.0% | 8.8% | 12.3% | 18.2% |
| Imaging-CT | 1.1% | 2.2% | 4.3% | 6.0% | 0.2% | 0.6% | 1.4% | 3.4% | 0.8% | 0.9% | 1.4% | 2.5% | 1.3% | 2.7% | 4.3% | 5.5% |
| Total Episode Cost - Median (Q1, Q3) |  |  |  |  |  |  |  |  |  |  |  |  |  |  |  |  |
| Total Episode Cost | \$106<br>(30,<br>220) | \$218<br>(97,<br>537) | \$455<br>(202,<br>1131) | \$835<br>(364,<br>1926) | \$165<br>(68,<br>397) | \$320<br>(149,<br>658) | \$573<br>(293,<br>1071) | \$877<br>(488,<br>1661) | \$528<br>(250,<br>1041) | \$688<br>(418,<br>1172) | \$1,012<br>(592,<br>1693) | \$1,333<br>(818,<br>2297) | \$229<br>(121,<br>597) | \$504<br>(236,<br>1205) | \$920<br>(429,<br>1867) | \$1,343<br>(719,<br>2473) |

**Figure 6.**
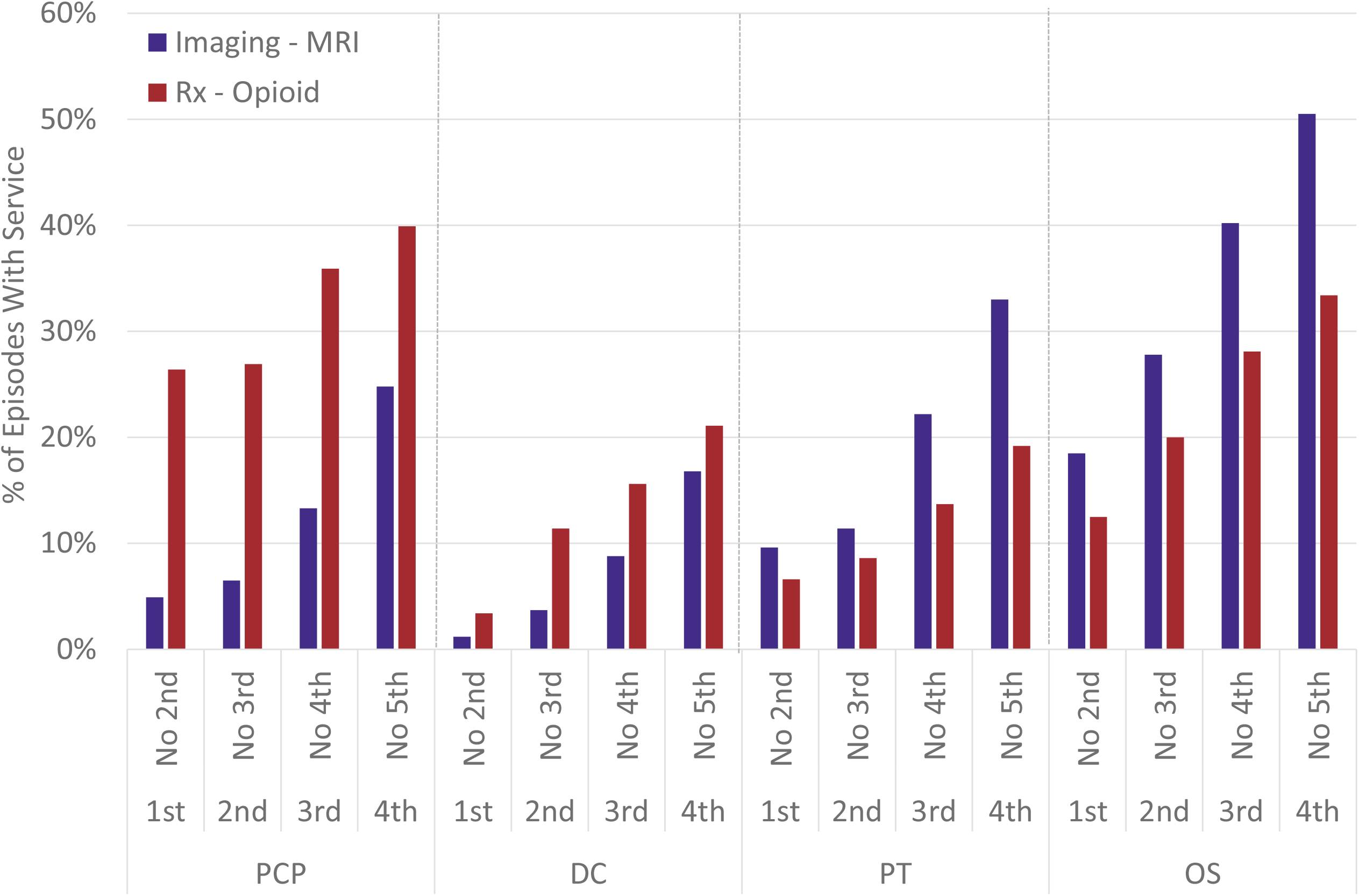
% of episodes with an MRI scan or prescription opioid for episodes where a primary care physician (PCP), chiropractor (DC), physical therapist (PT) or orthopedic surgeon (OS) was the last health care provider seen based on whether the PCP, DC, PT, or OS where the 1st, 2nd, 3rd or 4th HCP seen during an episode

**Figure 7.**
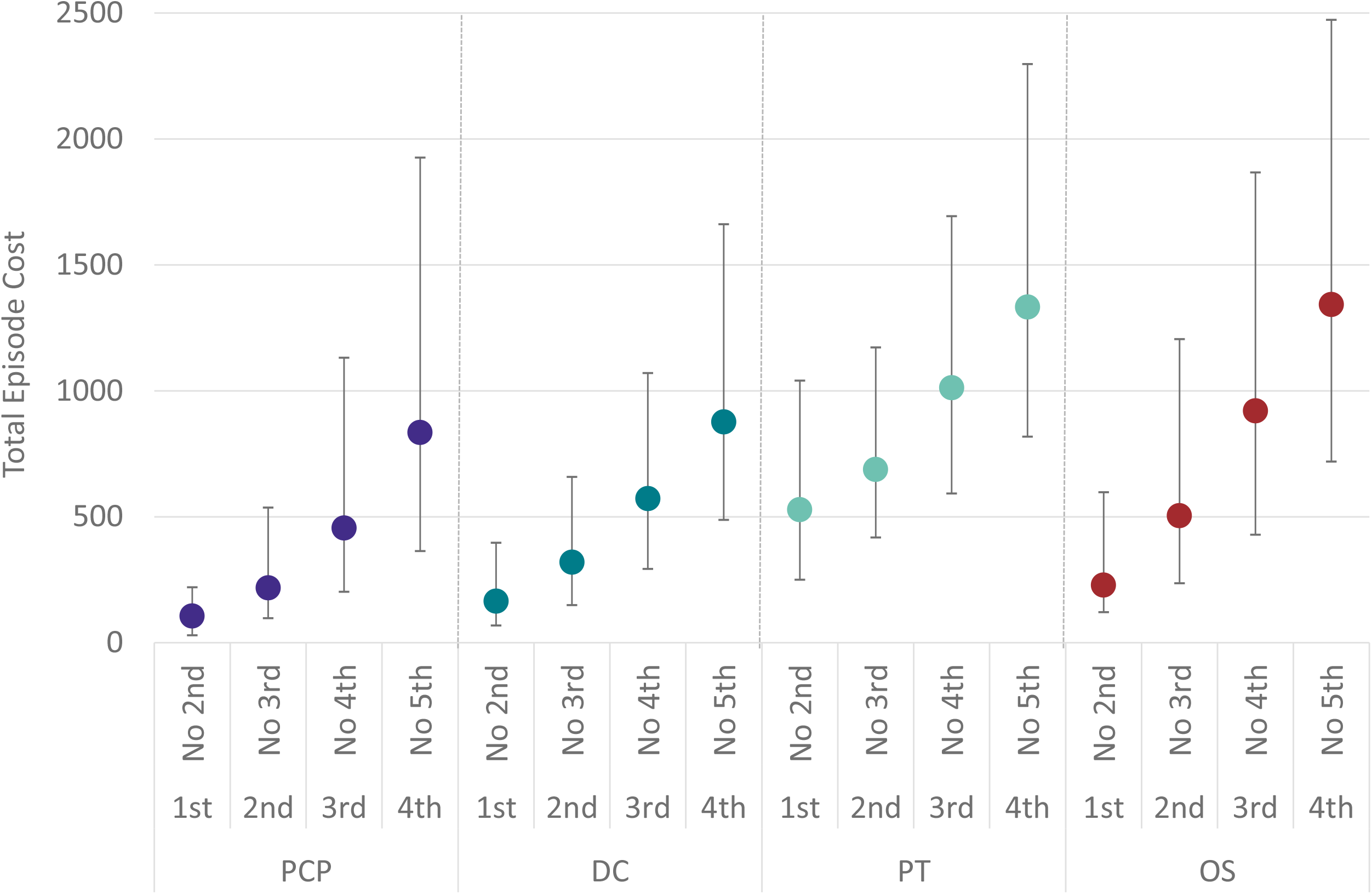
Median total episode cost and interquartile range (Q1, Q3) for episodes where a primary care physician (PCP), chiropractor (DC), physical therapist (PT) or orthopedic surgeon (OS) was the last health care provider seen based on whether the PCP, DC, PT, or OS where the 1st, 2nd, 3rd or 4th HCP seen during an episode

## Discussion

As a novel measure with many potential confounders and in the absence of clinical outcomes data it is important not to over interpret the finding that non-prescribing HCPs, particularly DCs and PTs, are most likely to be the last HCP seen for an episode of LBP. When coupled with the growing body of evidence demonstrating high levels of CPG concordance when these same types of HCP are initially contacted by individuals with LBP, additional exploration of attributes of the last HCP seen is warranted to more thoroughly understand the role these types of HCP play in managing LBP. While non-prescribing HCPs are most likely to be the last HCP seen for an episode of LBP, the rate of use of second- and third-line services and total episode cost increases the later non-prescribing HCPs are involved in an episode. This may provide additional support for positioning non-prescribing HCPs early in an episode of LBP as a strategy for improving CPG concordance and value.

This study has several limitations including its retrospective design and those associated with the use of administrative databases. The cohort had continuous highly uniform commercial insurance coverage and the processing of administrative claims data included extensive quality and actuarial control measures, nonetheless, data errors, variability in benefit plan design, variability in enrollee cost-sharing responsibility, and missing information were potential sources of confounding or bias. Although the commercial insurer HCP database is under continual validation it may have included errors or missing information. Summarizing total episode cost has potential limitations associated with insurance coverage, nature of network participation, and alternative reimbursement models. While individuals from all 50 states and most US territories were included, providing a measure of generalizability, the cohort did not describe a U.S representative sample.

Another important limitation was the risk of confounding and bias associated with the limited ability to control for individual preference for types of HCP seen during an episode of LBP, individual expectations, or requests for specific health care services, and potentially meaningful differences in clinical complexity of individuals contacting or being referred to different types of HCP. This limitation was partially addressed by narrowing the study population with several exclusions. The study excluded LBP associated with serious pathology, individuals with multiple episodes of LBP, and episodes involving a surgical procedure. Sub analyses were conducted to provide insight into to the degree to which individual and population factors may have influenced results.

While this study focused on individuals with a single episode of LBP, without the availability of clinical outcomes or patient self-report data, the last HCP seen cannot be assumed to be associated with, or a proxy for, resolution of LBP. Individuals may have been dissatisfied with care and discontinued all treatment, may have paid for additional treatment on a self-pay basis, or elected to pursue self-management. With the cohort having continuous insurance coverage during the study period and a median post-episode clean period of 401 days (Q1 248, Q3 638, minimum 61) during which there was no services provided for LBP, it is highly unlikely that after the last HCP seen there was a worsening of LBP symptoms. In future analyses, expanding the cohort to include individuals with multiple episodes of LBP will be important.

The volume of data involved in exploring the many associations in the 1.4 million possible HCP combinations presented another limitation. This will be addressed through subsequent analyses of the associations between HCP sequence patterns, cohort characteristics,rates of exposure to the types of health care services and total cost.

With the last HCP seen for LBP being a novel measure, it is challenging to corroborate previous studies. When involved in an episode of LBP as either the initial or subsequent HCP, the finding that DCs are more likely to be the last HCP seen, and associated with the lowest total cost, of any type of HCP is consistent with DCs emphasis on CPG concordant non-pharmaceutical and non-interventional services. ^3^ The finding that PTs, are more frequently contacted as a second HCP than as an initial HCP is consistent with PTs historically being positioned as referral recipients rather than initial contact HCPs. ^26,27^ When contacted as a second, third or fourth HCP PTs are as likely as a DC to be the last HCP seen which makes sense given the similar emphasis on CPG recommended first-line services. ^3^ The finding of PTs being infrequently contacted as the initial HCP is consistent with a previous study ^3^, however when contacted as the initial HCP, PTs are less likely than PCPs to be the last HCP seen which indicates the potential for less practice autonomy than DCs and warrants further study. A possible interpretation of these findings is that DCs and PTs possess similar knowledge, skills and training to effectively resolve LBP, with DCs positioned as portal of entry autonomous HCPs and PTs positioned as a referral recipient from and referral source to other types of HCPs.

## Conclusions

The last HCP seen by an individual with LBP is a novel area of investigation and further exploration is warranted to understand whether this study’s findings can be replicated and whether the approach provides meaningful insights into the types of HCP that are most helpful in resolving LBP. Whether seen initially, or later in an episode, DCs are most likely to be the last HCP seen by an individual with LBP. PTs are similar, except when they are the initial HCP contacted by an individual with LBP. While DCs and PTs are most likely to be the last HCP seen during an episode of LBP the later in an episode a DC or PT is seen associated beneficial attributes such as opioid avoidance are lost, and total episode cost increases. This adds additional support to the growing body of literature advocating for positioning these types of HCPs earlier in an episode of LBP.

## Supporting information

Supplement 1 - Cohort Attributes

Supplement 2 - Cohort Attributes By Type of Initial Healthcare Provider

Supplement 3 - Cohort and Population Attributes By Type of Last Healthcare Provider

Supplement 4 - Type of Last Healthcare Provider Seen By Population Deprivation and Race/Ethnicity

Supplement 4 - Risk Ratio For Type of Last Healthcare Provider Seen By Population Deprivation and Race/Ethnicity

Supplement - STROBE Checklist

## Data Availability

All data produced in the present work are contained in the manuscript

## Declarations

### Ethics approval and consent to participate

Because the data was de-identified or a Limited Data Set in compliance with the Health Insurance Portability and Accountability Act and customer requirements, the UnitedHealth Group Office of Human Research Affairs determined that this study was exempt from Institutional Review Board review.

### Consent for publication

Not applicable

### Availability of data and materials

The data are proprietary and are not available for public use but, under certain conditions, may be made available to editors and their approved auditors under a data-use agreement to confirm the findings of the current study.

### Authors’ contributions

Study conception and design; **DE.** Data acquisition; **DE, MZ.** Data analysis and interpretation; **DE, MZ.** Draft or substantially revise manuscript; **DE, MZ.**

## List of Abbreviations

LBP: Low back pain
US: United States
CPG: Clinical practice guideline
PCP: Primary care provider
DC: Doctor of Chiropractic
PT: Physical Therapist
LAc: Licensed acupuncturist
HCP: Health care provider
ADI: Area Deprivation Index
WTP: Willingness to pay
STROBE: Strengthening the Reporting of Observational Studies in Epidemiology
ETG^*®*^: Episode Treatment Group^*®*^
ERG^*®*^: Episode Risk Group^*®*^
ACP: American College of Physicians
RR: Risk ratio
OR: Odds ratio
CI: Confidence interval
CMT: Chiropractic manipulative treatment
AC: Active care
MT: Manual therapy

