## Supplement 1 - Cohort Attributes for "Low back pain care pathways – is the last provider seen more important than the first: A retrospective cohort study"

| Supplement 1 - Cohort Attributes | | |
|---|---|---|
| Individuals | | 503958 |
| Episodes | | 503958 |
| # Unique Health Care Providers (HCP) | | 196522 |
| Total Cost | | 387867096 |
| Individual attributes - % or Median (Q1, Q3) | | |
| Female | | 52.9% |
| Age | | 44 (34, 54) |
| ERG Risk Score | | 1.3 (0.6, 2.8) |
| Individual Home Address 5 Digit Zip Code Attributes - Median (Q1, Q3) | | |
| Population % Non-Hispanic White (NWH) | | 72.7% (51.7%, 85.5%) |
| Area Deprivation Index (ADI) | | 45 (28, 62) |
| Household Adjusted Gross Income (AGI) | | 64,389 (50,223, 90,352) |
| HCP per 1000 - DC | | 0.23 (0.09, 0.43) |
| HCP per 1000 - PT | | 0.17 (0.04, 0.41) |
| HCP per 1000 - LAc | | 0.00 (0.00, 0.03) |
| Episode Attributes - Median (Q1, Q3) (Minimum) | | |
| Clean Period - Before Initial Episode - Days | | 646 (431, 865) (91) |
| Clean Period - After Final Episode - Days | | 405 (248, 638) (61) |
| Episode Duration - Days | | 28 (1, 141) (1) |
| Episode Cost | | $204 (78, 647) |
| # of HCP Seen | | 2 (1, 3) |
| Type of HCP Initially Contacted - Episodes (%) | | |
| Primary Care | Primary care provider (PCP) | 177265 (35.2%) |
| | Nurse | 37857 (7.5%) |
| | Physician's Assistant (PA) | 25350 (5.0%) |
| | Doctor of Osteopathy (DO) | 469 (0.1%) |
| Non-Prescriber | Doctor of Chiropractic (DC) | 130520 (25.9%) |
| | Physical Therapist (PT) | 5114 (1.0%) |
| | Licensed Acupuncturist (LAc) | 2634 (0.5%) |
| Specialist | Orthopedic Surgeon (OS) | 27799 (5.5%) |
| | Physical Medicine & Rehabilitatino (PMR) | 12496 (2.5%) |
| | Pain Management (PM) | 9013 (1.8%) |
| | Neurosurgeon (NS) | 3058 (0.6%) |
| | Neurologist (Neuro) | 3917 (0.8%) |
| | Rheumatologist (Rheum) | 3354 (0.7%) |
| | MD - Other | 13877 (2.8%) |
| Emergency /Urgent | Emergency Medicine (EM) | 15760 (3.1%) |
| | Radiology (Rad) | 30393 (6.0%) |
| | Urgent Care (UC) | 5082 (1.0%) |
