## Supplement 2 - Cohort Attributes By Type of Initial Healthcare Provider for "Low back pain care pathways – is the last provider seen more important than the first: A retrospective cohort study"

| Supplement 2 - Cohort attributes by type of initial health care provider (HCP) contacted | | | | | | | | |
|---|---|---|---|---|---|---|---|---|
| % or Median (Q1, Q3) | | # Episodes (%) | Individual attributes | | | Individual home address zip code population attributes | | |
| | | | % Female | Age | ERG® Risk Score | Area Deprivation Index (ADI) | Household Adjusted Gross Income (AGI) | % non-Hispanic White (NHW) population |
| Primary Care | Primary care provider (PCP) | 177265 (35.2%) | 52.9% | 46 (36, 55) | 1.4 (0.7, 2.8) | 46 (29, 64) | 62,793 (48,913, 87,588) | 69.5% (47.4%, 84.0%) |
| | Nurse | 37857 (7.5%) | 55.8% | 44 (34, 54) | 1.4 (0.6, 2.7) | 55 (38, 69) | 56,665 (46,621, 74,594) | 76.3% (56.5%, 88.5%) |
| | Physician's Assistant (PA) | 25350 (5.0%) | 52.3% | 44 (34, 54) | 1.3 (0.6, 2.7) | 47 (32, 63) | 62,052 (49,516, 83,108) | 75.1% (55.6%, 86.6%) |
| | Doctor of Osteopathy (DO) | 469 (0.1%) | 63.8% | 45 (35, 55) | 1.5 (0.7, 2.7) | 41 (26, 60) | 71,892 (55,139, 106,885) | 76.4% (58.8%, 86.3%) |
| Non-Prescriber | Doctor of Chiropractic (DC) | 130520 (25.9%) | 51.5% | 40 (31, 50) | 1.0 (0.4, 2.0) | 43 (28, 60) | 67,675 (53,316, 93,242) | 76.8% (59.7%, 88.1%) |
| | Physical Therapist (PT) | 5114 (1.0%) | 59.0% | 44 (34, 54) | 1.6 (0.8, 3.2) | 32 (19, 51) | 78,426 (57,402, 114,853) | 72.5% (53.2%, 83.8%) |
| | Licensed Acupuncturist (LAc) | 2634 (0.5%) | 65.7% | 40 (33, 50) | 1.1 (0.5, 2.5) | 23 (12, 36) | 89,698 (64,422, 132,336) | 60.9% (40.4%, 76.4%) |
| Specialist | Orthopedic Surgeon (OS) | 27799 (5.5%) | 52.0% | 48 (36, 56) | 1.8 (0.8, 3.4) | 38 (21, 56) | 74,460 (54,706, 112,306) | 68.7% (49.0%, 82.1%) |
| | Physical Medicine & Rehabilitation (PMR) | 12496 (2.5%) | 52.7% | 48 (37, 56) | 2.0 (0.9, 3.7) | 35 (19, 55) | 77,360 (55,597, 117,460) | 69.7% (50.4%, 82.5%) |
| | Pain Management (PM) | 9013 (1.8%) | 53.6% | 50 (41, 57) | 2.9 (1.5, 5.0) | 46 (29, 62) | 64,795 (49,810, 92,276) | 67.8% (49.0%, 81.6%) |
| | Neurosurgeon (NS) | 3058 (0.6%) | 47.5% | 50 (39, 57) | 2.4 (1.1, 4.1) | 44 (27, 62) | 68,771 (51,468, 102,885) | 72.1% (53.1%, 84.6%) |
| | Neurologist (Neuro) | 3917 (0.8%) | 62.5% | 49 (39, 56) | 2.8 (1.5, 5.0) | 42 (24, 59) | 68,005 (51,450, 98,214) | 67.6% (46.9%, 82.2%) |
| | Rheumatologist (Rheum) | 3354 (0.7%) | 72.2% | 50 (40, 57) | 3.0 (1.7, 4.8) | 45 (27, 62) | 65,381 (50,307, 93,503) | 70.4% (48.7%, 84.2%) |
| | MD - Other | 13877 (2.8%) | 53.5% | 47 (37, 56) | 2.3 (1.1, 4.3) | 43 (26, 61) | 66,594 (50,771, 95,516) | 70.6% (49.4%, 83.7%) |
| Emergency/ Urgent | Emergency Medicine (EM) | 15760 (3.1%) | 50.0% | 40 (31, 51) | 1.3 (0.6, 2.6) | 51 (33, 67) | 58,123 (45,859, 79,092) | 67.8% (41.9%, 84.0%) |
| | Radiology (Rad) | 30393 (6.0%) | 53.3% | 45 (33, 55) | 1.7 (0.8, 3.5) | 48 (30, 65) | 61,547 (48,053, 86,376) | 73.0% (49.5%, 86.5%) |
| | Urgent Care (UC) | 5082 (1.0%) | 50.8% | 40 (31, 50) | 0.9 (0.4, 1.9) | 39 (24, 56) | 66,163 (50,378, 90,791) | 64.8% (41.8%, 81.4%) |
