## Supplement 3 - Cohort and Population Attributes By Type of Last Healthcare Provider for "Low back pain care pathways – is the last provider seen more important than the first: A retrospective cohort study"

| Supplement 3 - Non-surgical low back pain cohort and local population attributes associated with type of health care provider (HCP) seen last based on sequence of HCP involvement in episode | | | | | | | | | | | | |
|---|---|---|---|---|---|---|---|---|---|---|---|---|
| Type of HCP | 1st HCP | 2nd HCP | 3rd HCP | 4th HCP | 1st HCP | 2nd HCP | 3rd HCP | 4th HCP | 1st HCP | 2nd HCP | 3rd HCP | 4th HCP |
| | No 2nd | No 3rd | No 4th | No 5th | No 2nd | No 3rd | No 4th | No 5th | No 2nd | No 3rd | No 4th | No 5th |
| | Individual - % Female | | | | Individual - ERG® Risk Score | | | | Zip Code - Household Adjusted Gross Income (AGI) | | | |
| PCP | 52.4% | 55.2% | 55.9% | 55.1% | 1.3 (0.6, 2.5) | 1.5 (0.8, 3.0) | 1.9 (1.0, 3.6) | 2.3 (1.2, 4.1) | 62,512 (48,686, 87,314) | 62,491 (48,907, 85,596) | 61,283 (48,540, 82,124) | 60,173 (48,385, 80,286) |
| Nurse | 56.0% | 55.5% | 57.2% | 58.5% | 1.2 (0.6, 2.5) | 1.5 (0.7, 2.9) | 1.8 (1.0, 3.6) | 2.4 (1.3, 4.1) | 56,355 (46,461, 73,712) | 56,523 (46,496, 73,707) | 56,379 (46,671, 73,501) | 56,696 (46,094, 75,629) |
| PA | 52.6% | 52.1% | 53.3% | 53.4% | 1.2 (0.5, 2.3) | 1.4 (0.7, 2.7) | 1.8 (0.9, 3.3) | 2.1 (1.1, 3.7) | 61,660 (49,308, 82,671) | 61,308 (49,034, 81,246) | 59,945 (47,919, 80,256) | 63,498 (49,863, 84,877) |
| DO | 64.4% | 70.7% | 60.7% | 75.0% | 1.3 (0.6, 2.6) | 2.0 (1.2, 3.1) | 2.9 (1.5, 4.1) | 1.2 (0.5, 2.1) | 72,666 (55,400, 106,019) | 71,729 (53,876, 111,161) | 76,931 (56,952, 94,238) | 62,446 (61,032, 76,753) |
| DC | 50.8% | 53.9% | 54.1% | 56.1% | 0.9 (0.4, 1.9) | 1.3 (0.7, 2.6) | 1.5 (0.8, 2.9) | 1.8 (0.9, 3.4) | 67,808 (53,384, 93,628) | 66,189 (52,702, 90,643) | 67,396 (52,832, 92,216) | 64,795 (51,740, 87,129) |
| PT | 59.3% | 55.1% | 54.6% | 54.2% | 1.4 (0.7, 2.9) | 1.3 (0.6, 2.6) | 1.4 (0.7, 2.8) | 1.5 (0.8, 2.9) | 77,923 (56,796, 114,374) | 77,363 (56,565, 112,463) | 77,437 (56,507, 110,546) | 75,051 (55,229, 106,906) |
| LAc | 65.6% | 67.0% | 60.9% | 62.2% | 1.0 (0.4, 2.3) | 1.4 (0.8, 2.8) | 1.6 (0.7, 2.6) | 1.6 (0.8, 3.3) | 89,170 (64,425, 132,126) | 92,400 (65,275, 133,396) | 103,584 (72,731, 133,954) | 87,140 (61,539, 117,491) |
| OS | 52.0% | 50.2% | 50.0% | 49.7% | 1.6 (0.8, 3.1) | 1.9 (0.9, 3.4) | 2.2 (1.0, 3.8) | 2.4 (1.2, 4.0) | 75,175 (54,730, 114,194) | 70,468 (52,778, 105,839) | 67,684 (52,031, 101,999) | 68,815 (52,173, 102,279) |
| PMR | 52.2% | 50.5% | 52.4% | 50.5% | 1.8 (0.9, 3.4) | 2.0 (1.0, 3.7) | 2.4 (1.2, 4.1) | 2.5 (1.2, 4.1) | 77,799 (55,671, 116,417) | 74,595 (54,020, 110,982) | 71,570 (53,017, 108,318) | 69,658 (52,395, 106,095) |
| PM | 52.5% | 52.9% | 53.5% | 51.3% | 2.8 (1.4, 4.6) | 2.8 (1.5, 4.7) | 2.9 (1.5, 4.7) | 3.1 (1.6, 5.1) | 65,353 (49,810, 93,818) | 64,177 (50,352, 89,602) | 64,297 (49,834, 90,352) | 63,885 (50,307, 91,003) |
| NS | 45.6% | 49.3% | 43.1% | 42.6% | 2.0 (0.9, 3.7) | 2.3 (1.1, 3.9) | 2.3 (1.1, 3.8) | 2.5 (1.2, 4.3) | 69,405 (52,360, 106,258) | 65,122 (50,805, 92,490) | 64,230 (48,501, 96,013) | 62,207 (49,356, 85,000) |
| Neuro | 60.8% | 58.5% | 57.8% | 62.7% | 2.6 (1.4, 4.4) | 2.7 (1.3, 4.7) | 2.9 (1.5, 5.3) | 3.6 (1.9, 5.8) | 69,197 (51,676, 100,639) | 67,589 (50,985, 96,009) | 67,271 (50,396, 95,772) | 63,582 (49,790, 87,160) |
| Rheum | 71.1% | 74.2% | 75.9% | 71.7% | 2.8 (1.6, 4.5) | 3.0 (1.7, 5.0) | 3.5 (1.8, 5.3) | 3.6 (2.2, 5.3) | 64,795 (49,759, 94,171) | 66,193 (49,807, 92,224) | 66,252 (51,257, 94,773) | 62,960 (50,371, 87,782) |
| MD Oth | 52.8% | 52.7% | 52.0% | 52.4% | 2.1 (1.0, 3.9) | 2.2 (1.1, 3.9) | 2.6 (1.4, 4.5) | 3.0 (1.7, 5.5) | 67,084 (50,985, 95,516) | 66,500 (51,457, 95,152) | 68,210 (51,562, 95,833) | 64,694 (50,262, 95,147) |
| EM | 50.5% | 50.7% | 53.1% | 59.6% | 1.1 (0.5, 2.3) | 1.4 (0.6, 2.8) | 1.7 (0.8, 3.3) | 2.4 (1.3, 4.6) | 57,854 (45,436, 78,906) | 56,850 (45,624, 77,489) | 57,293 (45,833, 73,371) | 58,705 (46,269, 79,994) |
| Rad | 54.3% | 53.1% | 51.0% | 54.0% | 1.6 (0.7, 3.3) | 1.4 (0.7, 2.8) | 1.8 (0.9, 3.4) | 2.3 (1.1, 4.3) | 60,763 (47,286, 84,972) | 62,483 (49,279, 86,747) | 63,498 (50,108, 88,611) | 63,294 (49,337, 94,574) |
| UC | 51.5% | 47.6% | 49.7% | 50.0% | 0.8 (0.4, 1.8) | 0.8 (0.4, 1.8) | 1.2 (0.6, 2.1) | 1.7 (0.8, 3.2) | 65,540 (49,628, 89,971) | 68,063 (53,220, 90,050) | 71,479 (53,657, 101,311) | 64,970 (51,507, 84,166) |
| | Individual - Age | | | | Zip Code - Area Deprivation Index (ADI) | | | | Zip Code - % Non-Hispanic White | | | |
| PCP | 46 (35, 55) | 45 (35, 55) | 47 (37, 56) | 48 (38, 56) | 47 (29, 64) | 47 (30, 64) | 49 (32, 65) | 49 (32, 66) | 68.6% (46.1%, 83.4%) | 71.8% (49.5%, 85.1%) | 73.0% (50.1%, 86.0%) | 73.5% (51.6%, 86.8%) |
| Nurse | 43 (33, 53) | 44 (34, 53) | 46 (36, 54) | 46 (37, 56) | 55 (39, 69) | 55 (39, 69) | 54 (38, 69) | 55 (38, 70) | 76.0% (55.7%, 88.3%) | 76.2% (56.2%, 88.3%) | 76.1% (56.4%, 89.0%) | 77.4% (57.2%, 88.8%) |
| PA | 43 (33, 53) | 44 (34, 54) | 46 (35, 55) | 48 (38, 55) | 48 (32, 64) | 49 (33, 64) | 50 (34, 65) | 48 (31, 63) | 75.0% (55.3%, 86.6%) | 74.4% (54.0%, 86.3%) | 74.4% (55.5%, 87.3%) | 76.2% (58.5%, 87.9%) |
| DO | 44 (35, 53) | 45 (34, 52) | 53 (40, 58) | 44 (31, 57) | 41 (26, 58) | 43 (28, 63) | 41 (29, 48) | 54 (39, 62) | 76.1% (58.6%, 86.3%) | 69.1% (59.9%, 86.7%) | 75.1% (62.2%, 84.4%) | 74.3% (61.2%, 86.7%) |
| DC | 39 (30, 50) | 42 (33, 52) | 43 (33, 53) | 45 (35, 54) | 43 (27, 60) | 45 (29, 61) | 44 (28, 60) | 44 (29, 60) | 76.7% (59.4%, 88.0%) | 77.0% (60.0%, 88.2%) | 76.6% (59.9%, 88.2%) | 76.6% (60.5%, 87.9%) |
| PT | 43 (33, 54) | 44 (34, 55) | 45 (35, 55) | 46 (35, 55) | 33 (19, 52) | 34 (19, 51) | 34 (19, 52) | 36 (21, 55) | 71.4% (52.4%, 83.2%) | 71.4% (52.3%, 82.9%) | 72.4% (52.9%, 83.7%) | 73.6% (54.5%, 84.6%) |
| LAc | 39 (33, 50) | 42 (33, 51) | 42 (36, 51) | 45 (37, 50) | 23 (12, 36) | 21 (12, 36) | 19 (11, 32) | 24 (10, 39) | 61.3% (40.2%, 76.5%) | 63.4% (41.5%, 77.3%) | 63.6% (47.7%, 78.3%) | 56.6% (31.2%, 77.8%) |
| OS | 47 (35, 56) | 47 (36, 56) | 48 (37, 57) | 48 (38, 57) | 37 (21, 57) | 41 (23, 58) | 42 (25, 60) | 42 (25, 61) | 67.6% (48.0%, 81.4%) | 69.4% (49.3%, 82.2%) | 70.2% (49.6%, 83.9%) | 72.4% (52.4%, 84.6%) |
| PMR | 47 (37, 56) | 48 (38, 56) | 49 (39, 57) | 50 (40, 57) | 35 (19, 55) | 36 (21, 55) | 39 (22, 57) | 41 (23, 59) | 68.7% (49.5%, 81.9%) | 71.2% (51.2%, 83.4%) | 72.7% (53.4%, 84.3%) | 74.4% (52.9%, 86.5%) |
| PM | 50 (40, 57) | 50 (41, 57) | 50 (41, 57) | 52 (42, 58) | 46 (28, 62) | 47 (30, 62) | 46 (29, 62) | 47 (29, 63) | 67.1% (48.0%, 81.1%) | 68.3% (47.3%, 82.2%) | 70.4% (51.2%, 82.9%) | 70.8% (50.4%, 84.3%) |
| NS | 49 (39, 57) | 50 (39, 58) | 50 (40, 58) | 51 (41, 57) | 43 (26, 61) | 46 (29, 63) | 48 (27, 67) | 49 (30, 67) | 71.3% (52.3%, 84.3%) | 71.9% (52.1%, 84.7%) | 73.3% (53.1%, 86.3%) | 76.2% (58.6%, 87.4%) |
| Neuro | 48 (38, 56) | 47 (37, 56) | 49 (39, 57) | 48 (39, 56) | 41 (24, 59) | 43 (25, 60) | 42 (24, 60) | 44 (28, 61) | 66.0% (44.8%, 81.3%) | 67.1% (46.2%, 81.2%) | 67.4% (42.7%, 80.0%) | 70.8% (49.5%, 84.0%) |
| Rheum | 50 (41, 57) | 50 (40, 57) | 51 (41, 58) | 49 (40, 57) | 45 (27, 62) | 44 (29, 62) | 45 (27, 60) | 45 (28, 62) | 69.0% (46.1%, 83.4%) | 68.7% (47.3%, 84.2%) | 69.5% (53.4%, 84.5%) | 73.2% (48.0%, 85.8%) |
| MD Oth | 47 (36, 56) | 47 (36, 56) | 48 (38, 56) | 50 (40, 58) | 43 (26, 61) | 43 (27, 61) | 43 (26, 61) | 45 (27, 63) | 69.9% (48.6%, 83.2%) | 70.5% (49.9%, 84.3%) | 73.0% (53.7%, 86.1%) | 73.5% (53.4%, 85.8%) |
| EM | 39 (30, 50) | 41 (31, 52) | 43 (32, 52) | 43 (33, 52) | 51 (33, 67) | 52 (35, 67) | 51 (36, 66) | 51 (34, 67) | 66.5% (40.5%, 83.4%) | 69.6% (43.5%, 84.5%) | 70.5% (44.2%, 86.8%) | 71.3% (45.0%, 86.1%) |
| Rad | 44 (32, 55) | 45 (34, 55) | 46 (35, 55) | 48 (37, 56) | 48 (30, 66) | 47 (29, 64) | 46 (28, 63) | 47 (27, 64) | 71.2% (46.4%, 85.5%) | 74.3% (52.6%, 86.9%) | 75.5% (55.8%, 87.5%) | 76.1% (54.8%, 88.6%) |
| UC | 39 (30, 50) | 40 (31, 50) | 40 (32, 51) | 46 (37, 54) | 40 (25, 57) | 35 (22, 51) | 34 (21, 49) | 43 (30, 55) | 63.4% (40.2%, 80.8%) | 68.6% (48.3%, 83.6%) | 71.4% (51.6%, 85.2%) | 77.0% (55.9%, 87.7%) |

PCP=primary care provider, DO=doctor of osteopathy, DC=doctor of chiropractic, PT=physical therapist, LAc=licensed acupuncturist, OS=orthopedic surgeon, PMR=physical medicine & rebilitation, PM=pain medicine, NS=neurosurgeon, Neuro=neurologist, Rheum=rheumatologist, MD Oth = other medical physician, EM=emergency medicine, Rad=radiologist, UC=urgent care
