## Supplement 4 - Type of Last Healthcare Provider Seen By Population Deprivation and Race/Ethnicity for "Low back pain care pathways – is the last provider seen more important than the first: A retrospective cohort study"

Supplement 4 - Percent of episodes where type of health care provider (HCP) was the last HCP seen by episode sequence and population Area Deprivation Index (ADI) and percent of the population that is non-Hispanic white (% NHW)

| | Initial HCP - No 2nd | | | | | | 2nd HCP - No 3rd | | | | | |
|---|---|---|---|---|---|---|---|---|---|---|---|---|
| | ADI 75-100 % NHW 0-25 | | ADI 50-75 % NHW 50-75 | | ADI 0-25 % NHW 75-100 | | ADI 75-100 % NHW 0-25 | | ADI 50-75 % NHW 50-75 | | ADI 0-25 % NHW 75-100 | |
| | Episodes | *% No 2nd* | Episodes | *% No 2nd* | Episodes | *% No 2nd* | Episodes | *% No 3rd* | Episodes | *% No 3rd* | Episodes | *% No 3rd* |
| PCP | 5641 | *74.0%* | 17431 | *71.6%* | 14563 | *69.3%* | 1182 | *67.3%* | 3995 | *65.3%* | 3343 | *64.9%* |
| Nurse | 1267 | *64.4%* | 4286 | *64.0%* | 2350 | *56.2%* | 406 | *65.0%* | 1429 | *61.1%* | 844 | *59.4%* |
| PA | 793 | *60.8%* | 2743 | *57.3%* | 2301 | *56.9%* | 280 | *58.6%* | 991 | *57.0%* | 826 | *53.3%* |
| DO | 5 | *80.0%* | 46 | *73.9%* | 67 | *76.1%* | 1 | *100.0%* | 3 | *100.0%* | 9 | *55.6%* |
| DC | 1629 | *84.0%* | 11124 | *84.3%* | 13206 | *84.9%* | 269 | *73.6%* | 1646 | *72.4%* | 1762 | *73.7%* |
| PT | 62 | *71.0%* | 318 | *61.3%* | 824 | *59.5%* | 105 | *72.4%* | 511 | *72.4%* | 1127 | *69.5%* |
| LAc | 25 | *92.0%* | 88 | *78.4%* | 408 | *83.1%* | 2 | *50.0%* | 16 | *62.5%* | 70 | *64.3%* |
| OS | 504 | *64.5%* | 2326 | *62.4%* | 3576 | *61.7%* | 210 | *56.7%* | 925 | *57.0%* | 1284 | *54.2%* |
| PMR | 219 | *62.6%* | 903 | *63.9%* | 1807 | *62.1%* | 111 | *55.0%* | 422 | *59.0%* | 797 | *57.7%* |
| PM | 226 | *66.8%* | 988 | *61.3%* | 793 | *61.8%* | 117 | *53.0%* | 486 | *52.7%* | 394 | *54.1%* |
| NS | 72 | *51.4%* | 280 | *49.3%* | 335 | *54.0%* | 36 | *58.3%* | 214 | *52.3%* | 218 | *45.0%* |
| Neuro | 69 | *71.0%* | 361 | *55.1%* | 431 | *57.5%* | 43 | *72.1%* | 169 | *55.0%* | 164 | *57.3%* |
| Rheum | 81 | *58.0%* | 318 | *65.1%* | 311 | *61.7%* | 43 | *55.8%* | 144 | *52.1%* | 131 | *49.6%* |
| MD Oth | 333 | *54.4%* | 1345 | *54.5%* | 1361 | *54.1%* | 168 | *63.1%* | 630 | *57.8%* | 651 | *59.4%* |
| EM | 802 | *64.8%* | 1568 | *61.3%* | 1093 | *56.2%* | 221 | *67.4%* | 495 | *56.8%* | 358 | *50.0%* |
| Rad | 1055 | *54.1%* | 2851 | *50.2%* | 2837 | *46.6%* | 667 | *53.5%* | 2172 | *49.9%* | 2398 | *47.1%* |
| UC | 175 | *77.7%* | 391 | *61.9%* | 443 | *58.7%* | 30 | *73.3%* | 113 | *65.5%* | 147 | *63.9%* |
| | 3rd HCP - No 4th | | | | | | 4th HCP - No 5th | | | | | |
| | ADI 75-100 % NHW 0-25 | | ADI 50-75 % NHW 50-75 | | ADI 0-25 % NHW 75-100 | | ADI 75-100 % NHW 0-25 | | ADI 50-75 % NHW 50-75 | | ADI 0-25 % NHW 75-100 | |
| | Episodes | *% No 4th* | Episodes | *% No 4th* | Episodes | *% No 4th* | Episodes | *% No 5th* | Episodes | *% No 5th* | Episodes | *% No 5th* |
| PCP | 537 | *61.8%* | 1855 | *62.1%* | 1468 | *59.8%* | 234 | *62.0%* | 954 | *57.0%* | 704 | *53.3%* |
| Nurse | 191 | *62.3%* | 743 | *51.7%* | 412 | *54.6%* | 89 | *46.1%* | 416 | *49.8%* | 244 | *46.3%* |
| PA | 129 | *60.5%* | 524 | *49.8%* | 392 | *44.9%* | 63 | *61.9%* | 285 | *41.4%* | 239 | *48.5%* |
| DO | 0 | *N/A* | 8 | *62.5%* | 9 | *66.7%* | 0 | *N/A* | 2 | *100.0%* | 5 | *20.0%* |
| DC | 99 | *61.6%* | 799 | *67.8%* | 801 | *67.5%* | 58 | *60.3%* | 357 | *63.6%* | 364 | *63.2%* |
| PT | 76 | *65.8%* | 369 | *66.1%* | 776 | *63.0%* | 49 | *71.4%* | 239 | *63.2%* | 461 | *62.0%* |
| LAc | 1 | *0.0%* | 6 | *50.0%* | 43 | *72.1%* | 2 | *100.0%* | 5 | *60.0%* | 18 | *55.6%* |
| OS | 142 | *62.0%* | 590 | *50.5%* | 741 | *52.6%* | 93 | *52.7%* | 348 | *51.1%* | 428 | *51.2%* |
| PMR | 90 | *56.7%* | 336 | *60.4%* | 545 | *61.5%* | 64 | *56.2%* | 219 | *56.2%* | 360 | *59.2%* |
| PM | 101 | *61.4%* | 433 | *55.9%* | 345 | *56.2%* | 72 | *44.4%* | 335 | *59.4%* | 249 | *53.0%* |
| NS | 40 | *52.5%* | 164 | *47.0%* | 198 | *42.9%* | 29 | *34.5%* | 124 | *42.7%* | 127 | *42.5%* |
| Neuro | 18 | *66.7%* | 101 | *57.4%* | 114 | *50.0%* | 12 | *41.7%* | 77 | *49.4%* | 60 | *41.7%* |
| Rheum | 25 | *60.0%* | 84 | *52.4%* | 74 | *54.1%* | 13 | *46.2%* | 63 | *49.2%* | 48 | *60.4%* |
| MD Oth | 89 | *58.4%* | 398 | *57.0%* | 422 | *52.1%* | 67 | *58.2%* | 287 | *48.1%* | 274 | *54.0%* |
| EM | 71 | *52.1%* | 166 | *48.8%* | 108 | *44.4%* | 35 | *57.1%* | 84 | *56.0%* | 51 | *43.1%* |
| Rad | 245 | *42.0%* | 969 | *44.9%* | 1088 | *40.9%* | 118 | *37.3%* | 498 | *36.1%* | 586 | *40.3%* |
| UC | 8 | *25.0%* | 25 | *44.0%* | 29 | *62.1%* | 3 | *33.3%* | 8 | *37.5%* | 11 | *27.3%* |

Cells in red have risk ratio 95% confidence interval crossing the PCP reference of 1

Cells highlighted with bold font have risk ratio 95% plus confidence interval greater than the PCP reference of 1 indicating greater likelihood to be the last HCP seen

PCP=primary care provider, DO=doctor of osteopathy, DC=doctor of chiropractic, PT=physical therapist, LAc=licensed acupuncturist, OS=orthopedic surgeon, PMR=physical medicine & rebilitation, PM=pain medicine, NS=neurosurgeon, Neuro=neurologist, Rheum=rheumatologist, MD Oth = other medical physician, EM=emergency medicine, Rad=radiologist, UC=urgent care
